# Diffusivity anisotropy signature of the slowly expanding lesions predicts progression independent of relapse activity in multiple sclerosis

**DOI:** 10.1101/2025.07.25.25332073

**Authors:** A Calvi, Pascual F Vivó, E Solana, E Lopez-Soley, B Kanber, S Alba-Arbalat, M Sepulveda, E Martinez-Hernández, JM Cabrera-Maqueda, E Fonseca, S Medrano-Martorell, A Saiz, Y Blanco, P Villoslada, Carrasco F Prados, E Martinez-Heras, S Llufriu

**Affiliations:** Neuroimmunology and Multiple Sclerosis Unit and Laboratory of Advanced Imaging in Neuroimmunological Diseases (ImaginEM), Hospital Clinic Barcelona, IDIBAPS, Universitat de Barcelona, Barcelona, Spain; Institute of Neurology, Faculty of Brain Sciences, University College London (UCL), United Kingdom; Centre for Medical Image Computing (CMIC), Department of Medical Physics and Biomedical Engineering, University College London, London, United Kingdom; National Institute for Health Research (NIHR), University College London Hospitals (UCLH), Biomedical Research Centre, United Kingdom; Neuroradiology Department, Centre de Diagnòstic per la Imatge (CDI). Hospital Clinic Barcelona, IDIBAPS, Universitat de Barcelona, Barcelona, Spain; Department of Neurology, Hospital del Mar, Pompeu Fabra University, Barcelona, Spain; e-Health Centre, Universitat Oberta de Catalunya, Barcelona, Spain

## Abstract

**Background:** Slowly Expanding Lesions (SELs) in Multiple Sclerosis (MS) are markers of chronic active lesions and seem to trigger disability. This study aimed to analyse spatial features of SELs through diffusion MRI and their clinical impact on progression independent of relapse activity (PIRA).

**Methods:** An observational study of MS subjects prospectively followed since 2011; inclusion required at least three longitudinal T1/T2-weighted and diffusion-weighted MRIs. Subjects followed clinical assessments, using Multiple Sclerosis Functional Composite (MSFC) and Expanded Disability Status Scale (EDSS). At MRI, lesions were categorised using non-linear deformation as definite or possible SELs, and non-SELs. Fractional anisotropy (FA) was extracted from each lesion core and perilesional area. Differences in FA values across core and perilesional areas by SEL category were assessed using the Mann-Whitney test. Associations with PIRA and clinical outcomes were evaluated using mixed-effects, logistic, and Cox regression models.

**Results:** 130 subjects underwent MRI (median 25 months) and clinical assessments (median follow-up 9.2 years), of which 29 (22%) developed PIRA. Of 4811 lesions, 8% were definite SELs. Definite SELs exhibited FA decline over time in core and perilesional areas compared to other lesions. Longitudinal core FA reductions within definite SELs associated with worse MSFC z-score evolution (β=0.03, 95%CI 0.01-0.05, p=0.003), higher odds for PIRA (OR=0.01, 95% CI 0.01-0.12, p=0.001), and predicted faster time to reach first PIRA event (HR=0.03 95% CI 0-0.49, p=0.015).

**Conclusions:** Definite SELs show distinct greater microstructural damage and are associated with PIRA, making their FA signature a potential predictor of MS progression.

**What is already known on this topic:** early detection of neuroaxonal damage in chronic active multiple sclerosis (MS) lesions is highly relevant for evaluating progression independent of relapse. MRI can identify such damage by detecting slowly expanding lesions (SELs) and assessing spatial diffusivity measures. Analysing damage within the core and perilesional areas of these lesions and their relationship to disability may clarify the mechanisms driving MS progression.

**What this study adds:** This research demonstrates that diffusion MRI reveals distinct microstructural damage in chronic active MS lesions, providing valuable markers for predicting disease progression. By examining fractional anisotropy (FA) within the core and perilesional regions of SELs, a distinctive diffusion pattern of microstructural damage was identified. This SEL-specific FA reduction correlates with higher odds and earlier onset of progression in an MS cohort assessed shortly after diagnosis.

**How this study might affect research and clinical practice:** This study underscores the potential of FA reductions in SELs as early indicators of MS progression. Incorporating diffusion MRI, to assess lesion evolution into clinical practice could enhance the early detection of subclinical disease progression, enabling timely therapeutic interventions. Implementing these techniques may improve MS management protocols and patient outcomes.

## Introduction

Disability in multiple sclerosis (MS) is predominantly influenced by incomplete relapse recovery and progression independent of relapse (PIRA). PIRA can manifest from early stages in people with multiple sclerosis (pwMS) and it is thought to be linked to compartmentalised chronic inflammation^1^. The complexity of chronic inflammation is not fully understood but appears to result from various interconnected factors: accumulation of chronic active lesions in the white matter^2^, subpial and grey matter cortical lesions^3^, and leptomeningeal infiltrates^4^. Persistent low-grade inflammation in chronic active lesions seems to be linked to long-term disability in pwMS^5^. Those lesion types exhibit organised inflammation causing tissue damage at the core and activated macrophages-microglia at the lesion edge^6^, with progressive disease and a shorter time to disability accumulation^7^. Accumulation of chronic active lesions may substantially impact clinical disability progression and monitoring these lesions *in vivo* could indicate the risk for earlier PIRA events.

Recent consensus^8^ highlights the importance of identifying Paramagnetic Rim Lesions (PRLs) and Slowly Expanding Lesions (SELs) as MRI markers for chronic active lesions in MS. PRLs detect iron-laden deposits with high sensitivity through susceptibility-weighted imaging supported by imaging-pathological correlations^9^. Previous evidence demonstrated that PRLs correlate with MS disability^10^ and other markers of progression, such as spinal cord atrophy^11^. Despite this evidence, PRL detection on clinical MRI is still limited due to challenges with visual rim identification. Further studies in MS show that SELs are more prevalent than PRLs^12^ and SEL volume has been associated with disability^13^, highlighting the potential as a clinically relevant biomarker. SEL computation involves automated registration of longitudinal T1-weighted sequences and extraction of deformable voxels to track focal volume changes over time^14^. Notably, quantitative MRI methods revealed structural damage in chronic active lesions, such as increased T1 hypointensity within SELs^15^, indicating progressive neuroaxonal damage.

Diffusion tensor imaging (DTI) changes in MS tissues more specifically correlate with myelin content and axonal count, indicating neurodegeneration^16^. DTI-derived metrics, such as fractional anisotropy (FA), have been further explored in MS tissues helping to classify lesion types with varying microstructural damage, linked to clinical and cognitive decline^17^. A recent study found that lesions co-localized with both SEL and PRL labels had lower FA compared to the rest of T2 lesions, showing greater DTI damage through a decreased FA over time^18^. However, the characteristics of microstructural changes in the core and perilesional areas in SELs, and the clinical predictive value of their diffusivity signature remains unclear.

This study aimed to characterise chronic active lesions using FA, as it represents a sensitive DTI metric of axonal loss and demyelination, and to investigate the correlation of anisotropy within SELs and disability in MS. We hypothesised that SELs would exhibit greater microstructural damage, reflected in distinct diffusion anisotropy patterns, and be linked to PIRA. The specific aims of our study were: (1) to assess FA values in relevant spatial localisations (lesion core and perilesional area) cross-sectionally and longitudinally in SELs, and to compare them with the characteristics of other lesions; (2) to explore associations between FA values and MRI markers of disease progression (i.e. brain atrophy); and (3) to evaluate the predictive value of FA within SELs for disability outcomes and PIRA in pwMS.

## Materials and Methods

### Inclusion criteria & clinical assessments

This study analyses prospectively collected data from an observational cohort of pwMS^19^ enrolled since January 1st 2011, following STROBE guideline^20^. Ethical approval was granted by the local ethics committee (Comité de Ética de la Investigación con medicamentos del Hospital Clínic de Barcelona, registred act HCB/2022/0973), and all participants provided informed consent. Inclusion criteria were: (1) MS diagnosis (McDonald 2010 criteria), (2) at least three MRI sessions including conventional and diffusion-weighted imaging (DWI) within a maximum of 5 years, and (3) long-term clinical assessments. Of the 171 pwMS cohort, 130 were included in the study; 41 were excluded due to incomplete MRI sessions or DWI artefacts. Baseline data included age, gender, disease duration, and MS phenotype: relapsing-remitting (RRMS), primary progressive (PPMS) or secondary progressive (SPMS).

Clinical evaluations included the Expanded Disability Status Scale (EDSS)^21^ and Multiple Sclerosis Functional Composite (MSFC)^22^, consisting of the Nine Hole Peg Test (9HPT), Timed 25 Foot Walk (T25FWT), and Paced Auditory Serial Addition Test (PASAT) – 3 seconds per digit version converted to standardised z-scores. Neuropsychological assessments also included the Symbol Digit Modalities Test (SDMT) adjusted for age and education (Spanish normative data)^23^. Cognitive worsening was defined as a decrease of ≥20% in both the PASAT and the SDMT z-scores.

PIRA was assessed via EDSS scores^24^ at each visit. Baseline EDSS was recorded at the first visit, and, in case of a relapse, a re-baseline EDSS was taken three months post-relapse. PIRA event was defined as confirmed disability accumulation unrelated to EDSS changes within the relapse window, and characterised by an EDSS increase of 1.5, 1.0 to 4.5, or 0.5 if the baseline/re-baseline EDSS was 0, 1.0, or ≥5.0, respectively. The PIRA event was confirmed through the last follow-up visit, and only the first PIRA event was analysed. PwMS who did not develop PIRA over the final visit were defined as “Stable” as opposed to the “PIRA” group.

### MRI acquisitions and diffusion processing

In a longitudinal observational study involving 130 subjects, 390 MR images were acquired across multiple time points using 3T Siemens Magnetom Trio/Prisma fit scanners with 32/64-channel head coils, employing three different DWI acquisition protocols (Supplementary Table 1). Structural sequences included a 3D-Magnetization Prepared Rapid Acquisition Gradient Echo (MPRAGE) and 3D-T2 Fluid-attenuated inversion recovery (FLAIR). DWI preprocessing included denoising, Gibbs ringing removal, motion correction, B-matrix transformation, geometric unwarping by gradient field maps or TOPUP method, and bias field correction. FA was calculated using FSL (DTIFIT toolbox) with b-values of 1000–1500 s/mm², employing a weighted least square fitting method^25^. Focusing on a single FA metric was justified to simplify data interpretation and minimise the risk of type I errors associated with multiple testing. Scanner protocol variability across the entire dataset was minimised using ComBat harmonisation^26^(Supplementary Figure 1).

### Lesion segmentation and perilesional area definition

Lesion masks at baseline were extracted from MPRAGE and FLAIR using a 3D convolutional neural network (nnU-Net; https://github.com/MIC-DKFZ/nnUNet)^27^, and new lesions were identified by comparing FLAIR images from baseline to the last available follow-up, also using nnU-Net architecture^28^. Quality control for segmentation accuracy and new lesion detection was performed by an experienced neurologist (AC), excluding lesions smaller than 27 mm³ as previously described^29^. The perilesional area corresponded to 1 concentric voxel ring surrounding the outer part of the lesion mask. Lesion-filled T1w images were generated^30^ for determining cross-sectional normalised tissue types using the FSL-SIENAX tool^31^: normalised grey matter (NGM), normal-appearing white matter (NAWM), normalised lesion volume (NLV) and normalised brain volume (NBV). Percentage brain volume change (PBVC) from baseline to the last available follow-up MRI scan was calculated using SIENA^32^ and was annualised.

### SEL computation and diffusion anisotropy analysis

To identify SELs, Jacobian expansion values were computed to quantify localised volumetric changes within lesions over time. Deformation maps were generated via non-linear registration of longitudinal T1-weighted scans using “NiftyReg” (https://github.com/KCL-BMEIS/NiftyReg). The determinant of the Jacobian, derived from these deformation fields, served as a measure of lesion volume change and was used to classify three lesion types^15^: (1) definite SELs characterised by either constancy and concentricity of their expansion (Jacobian increasing at each time point and in concentric sub-lesion bands from the centre to the periphery); (2) possible SELs (overall positive Jacobian expansion); and (3) non-SELs as the rest of non-expanding lesions. The baseline counts and volumes of each lesion type were quantified globally and per subject. Finally, FA maps were registered to the T1-weighted space using boundary-based rigid registration, enabling the computation of the diffusion anisotropy values for different lesion types at relevant spatial localisations (lesion core and perilesional area) across all time points.

### Statistical analysis

The analysis was conducted using R (R Core Team [2020]) and Python 3.10 with open-source libraries, and statistical significance was set at p<0.05. Descriptive statistics were calculated for demographic, clinical, and MRI variables. Normality was assessed using the Shapiro-Wilk test and visual inspection. Data were presented as median (interquartile range, IQR) or mean (standard deviation, SD). As appropriate, group comparisons between PIRA and Stable patients were performed using Wilcoxon signed-rank, Kruskal-Wallis, or univariate linear regressions. Correction for multiple comparisons used Benjamini-Hochberg adjustment.

Lesion-level FA was analysed cross-sectionally and plotted using bootstrap technique (baseline and last follow-up MRI), examining the core and perilesional area across lesion types. Differences in lesion FA between SEL types at baseline and follow-up were analysed using the Mann-Whitney U test. The FA change was determined by calculating the difference between the baseline and last MRI measures for each lesion, and the mean FA change was grouped according to the lesion types. Mixed-effects models evaluated longitudinal change in core and perilesional FA in each lesion type, adjusted for scanner protocol. Correlations (Spearman) were explored between mean baseline FA and mean FA change in SELs and NLV or PBVC, adjusting for age and gender. Mixed-effects models evaluated all the clinical variables as the outcome and the interaction term of core and perilesional FA with time across lesion type as the predictor, including as fixed effects the demographic variables (age, gender), baseline lesion volume, PBVC and as random effects the lesion and the subject-level identifiers.

Logistic and Cox regression models were used to examine the risk of PIRA and to reach the first PIRA event. We investigated the subject-level mean FA (across all MRI time points) by lesion type within both core and perilesional areas and the odds of PIRA. These models were adjusted for age, gender, lesion count, Jacobian expansion, baseline lesion volume (specific to each lesion type), PBVC, and baseline EDSS, reporting odds ratios (OR) or hazard ratios (HR) and their 95% confidence interval (CI).

### Data availability

Imaging data in BIDS format and the clinical data supporting the findings of this research can be accessed upon reasonable request to the corresponding authors.

## Results

### Clinical, MRI characteristics and comparisons by PIRA group

Among 130 pwMS (Table 1: whole cohort), 93 were females (72%), the median age was 41.5 years old (IQR 34.6, 49.2), and disease duration at study onset was 6.8 (IQR 2.8, 13.5). The majority of them were diagnosed with relapsing MS (118, 91%) and 97 (75%) received a disease-modifying treatment (DMT), showing a baseline median EDSS of 1.5 (IQR 1.0, 2.0). Clinical assessments reached a median follow-up of 9.2 years (IQR 5.1, 10.1). Patients presented a median of 2 relapses (IQR 2, 6) during the study time. Regarding cognitive function, 15% of pwMS (20 out of 130) exhibited a decline in both SDMT and PASAT z-scores.

**Table 1.** Demographic, clinical and MRI and global diffusivity measures in the PIRA and stable groups.

|  | **Whole cohort**<br>**(n=130)** | **PIRA**<br>**(n=29)** | **Stable**<br>**(n=101)** | **P-value\*** |
| --- | --- | --- | --- | --- |
| Female, n (%) | 93 (72 %) | 24 (83%) | 69 (68%) | 0.136 |
| Disease duration, median (IQR) years | 6.8 (2.8, 13.5) | 11.4 (6.2, 15.7) | 5.7 (2.5, 12.5) | **0.016** |
| Age, median (IQR) years | 41.5 (34.6, 49.2) | 47.2 (43.5, 52.6) | 38.7 (33.1, 46.2) | **0.001** |
| Number of patients treated with any DMT at the last visit (%) | 97 (74%) | 19 (66%) | 78 (77%) | 0.342 |
| **Clinical** |  |  |  |  |
| EDSS baseline, median (IQR) | 1.5 (1.0, 2.0) | 2.5 (1.5, 4.5) | 1.5 (1.0, 2.0) | **<0.001** |
| EDSS change, median (IQR) | 0.5 (0, 1.5) | 1.5 (0.5, 4.5) | 0.0 (-1.0, 3.0) | **<0.001** |
| MSFC z-score baseline, mean (SD) | 0.2 (0.5) | 0.1 (0.5) | 0.3 (0.5) | 0.134 |
| 9HPT z-score baseline, mean (SD) | 0.3 (0.7) | 0.2 (1.0) | 0.4 (0.6) | 0.531 |
| T25FW z-score baseline, mean (SD) | 0.4 (0.2) | 0.4 (0.3) | 0.5 (0.2) | **0.001** |
| PASAT z-score baseline, mean (SD) | -0.2 (1.2) | -0.3 (1.4) | -0.2 (1.2) | 0.725 |
| SDMT z-score baseline, mean (SD) | 0.3 (1.1) | 0.1 (1.1) | 0.4 (1.0) | 0.361 |
| **MRI measures** |  |  |  |  |
| NBV, mean (SD), ml | 1465.1 (79.2) | 1437.2 (89.0) | 1473.0 (74.7) | 0.055 |
| NGM volume, mean (SD), ml | 747.2 (50.7) | 731.4 (53.1) | 751.8 (49.2) | 0.070 |
| NAWM volume, mean (SD), ml | 718.2 (45.2) | 705.9 (51.7) | 721.3 (42.8) | 0.151 |
| NLV, mean (SD), ml | 9.7 (12.1) | 12.7 (13.8) | 8.8 (11.5) | 0.063 |
| PBVC (annualised), median (IQR) (%) | -0.37 (-0.63, -0.16) | -0.61 (-0.90, -0.31) | -0.36 (-0.54, -0.14) | **0.002** |
| Baseline lesion count, median (IQR) | 31 (18, 47) | 34 (27, 55) | 30 (17, 45) | 0.072 |
| New lesion count (at last MRI ), median (IQR) | 1 (0, 4) | 1 (0, 4) | 0 (0, 4) | 0.565 |
| New lesion volume (at last MRI ), mean (SD), ml | 1.0 (4.2) | 1.7 (7.4) | 0.8 (2.6) | 0.666 |
| Lesion count, median (IQR) |  |  |  |  |
| Non-SELs | 19 (10, 31) | 24 (17, 35) | 18 (10, 30) | 0.070 |
| Possible SELs | 8 (4, 15) | 11 (5, 16) | 8 (4, 14) | 0.229 |
| Definite SELs | 2 (1, 5) | 2 (1, 4) | 2 (0, 5) | 0.431 |
| Baseline lesion volumes, mean (SD), ml | 4.5 (7.2) | 5.8 (8.2) | 4.2 (6.9) | 0.141 |
| Non-SELS | 1.2 (1.8) | 1.5 (1.4) | 1.1 (1.9) | **0.048\*\*** |
| Possible SELs | 1.3 (2.9) | 2.0 (4.0) | 1.1 (2.4) | 0.266 |
| Definite SELs |  |  |  |  |
| Jacobian expansion, mean (SD), (%) |  |  |  |  |
| Non-SELS | -5.9 (6.3) | -8.2 (9.9) | -5.5 (5.4) | **<0.001** |
| Possible SELs | 3.2 (3.2) | 5.43 (6.8) | 2.87 (2) | **<0.001** |
| Definite SELs | 6.3 (6.3) | 7.13 (8.3) | 6.19 (6) | 0.46 |
| **MRI diffusivity (baseline)** |  |  |  |  |
| FA in NAWM, mean (SD) | 0.59 (0.14) | 0.57 (0.15) | 0.60 (0.13) | 0.089 |
| FA in Lesion Core, mean (SD) | 0.38 (0.05) | 0.37 (0.04) | 0.38 (0.05) | 0.323 |
| FA in Perilesional Area, mean (SD) | 0.43 (0.05) | 0.43 (0.05) | 0.43 (0.05) | 0.688 |

\*PIRA and Stable groups were tested and the corresponding p-value was reported using the Mann–Whitney U test for the continuous variables and the $\chi$ -squared test for the categorical variables. Variables correspond to the baseline time point except for those indicated as change. \*\*P-value was not significant after B-H adjustment.
EDSS change corresponds to the difference between the final visit and baseline EDSS.
Abbreviations: PIRA = progression independent of relapse activity, EDSS = expanded disability status scale, Nine Hole Peg Test = 9HPT, Timed 25 Foot Walk = T25FW, Symbol Digit Modalities Test = SDMT, Paced Auditory Serial Addition Test = PASAT; Multiple Sclerosis Functional Composite = MSFC, gray matter = GM, normal-appearing white matter = NAWM, normalized brain volume = NBV, percentage brain volume change = PBVC, fractional anisotropy = FA.

The MRI analysis and processing were conducted in three longitudinal acquisitions over a median interval time of 25 months (IQR 24, 27). The median number of new lesions at the last MRI session was 1 (IQR 0, 4). Out of 4811 lesions analysed from the baseline, 400 were classified as definite SELs (8%), 1391 as possible SELs (29%), and 3020 were non-SELs (63%). At the patient level (Table 1), the median baseline lesion count was 31 (IQR 18, 46), corresponding to a median of 2 definite SELs (IQR 1, 5), 8 possible SELs (IQR 4, 15), and 18 non-SELs (IQR 10, 31). The annualised median PBVC from baseline to the last follow-up was −0.37% (IQR −0.63, −0.16).

In this cohort 29 pwMS (22%) developed PIRA by the final assessment. As shown in Table 1, the PIRA group had longer disease duration, higher EDSS (baseline and change), and older age. The proportions of patients on any DMT by the last follow-up did not differ between the groups. PwMS who developed PIRA were also characterised by greater PBVC reduction (−0.61% vs. −0.35%, p=0.001), with trends toward lower NBV and higher possible SEL volume (not significant after multiple comparisons adjustment). No baseline differences were found in FA values within NAWM, lesion core, or perilesional area between the two groups.

### Cross-sectional and longitudinal FA changes in lesion types

Figure 1 illustrates a pwMS with two lesions classified as definite SELs, identified through constant and concentric expansion, and the effect on microstructural integrity, reflected by areas of FA reduction. Normalised mean FA values at baseline and last follow-up were plotted for the core and the perilesional areas across lesion types (Figure 2). Definite SELs consistently showed the lowest core FA values at baseline and last follow-up compared to possible SELs and non-SELs (p<0.001, Kruskal-Wallis tests after BH adjustment). No cross-sectional differences were identified in the perilesional FA (baseline and last follow-up) between the definite SELs and the other lesion types.

**Figure 1.**
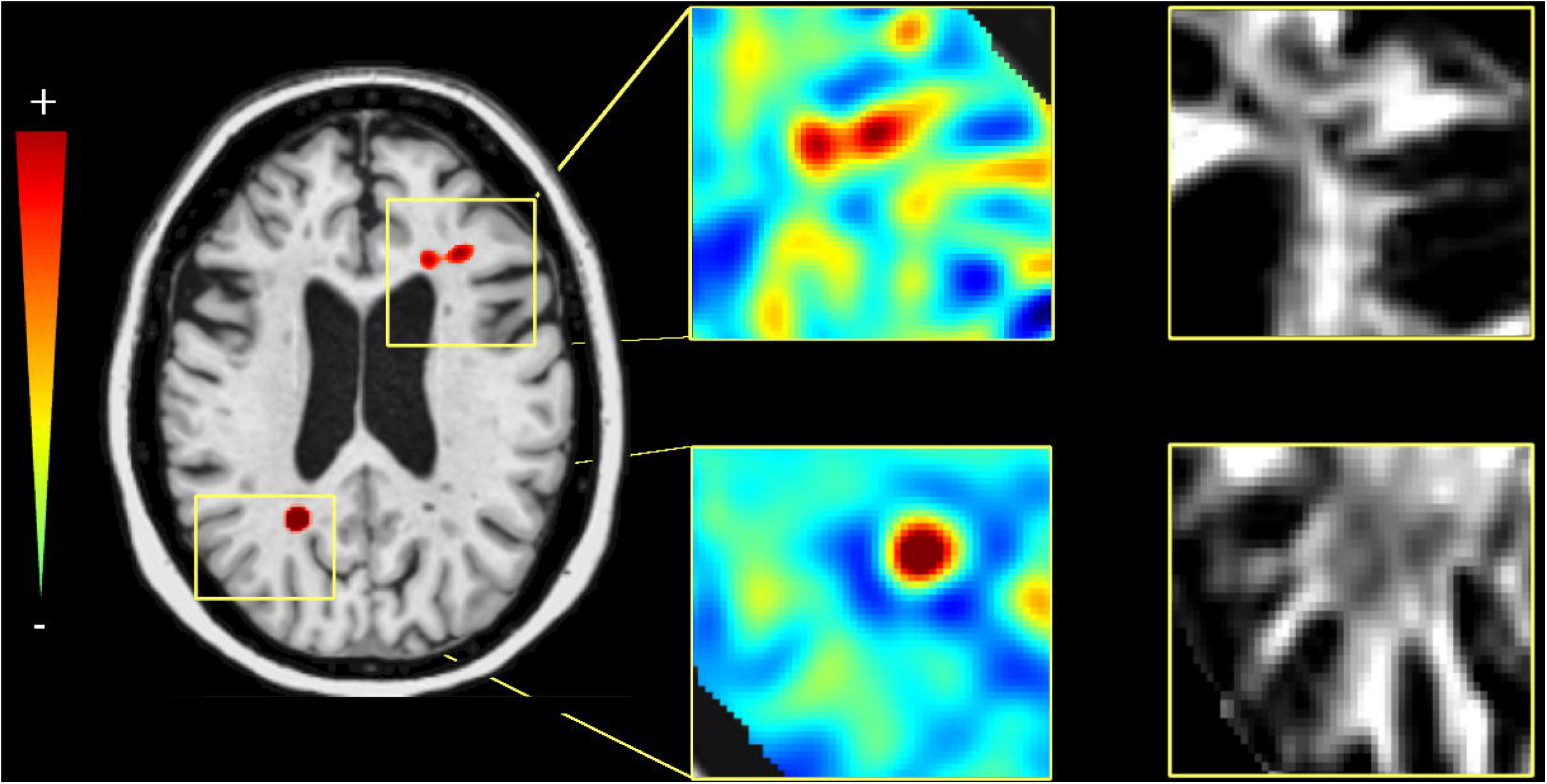
“Characterization of SELs and their association with decreased FA measures”. On the left side, an example MRI scan from a participant (patient in their 50s, EDSS 3.5 at baseline, total lesion volume 16.6 ml) as observed on MPRAGE axial view with the superimposed relative lesion mask. In the centre, two examples of definite SELs are magnified. The magnified left colour images show these SELs on their respective Jacobian maps, where red indicates higher values, suggesting volume expansion. The magnified right images display the corresponding DWI with FA maps, where hypointensity indicates higher tissue integrity disruption.

**Figure 2.**
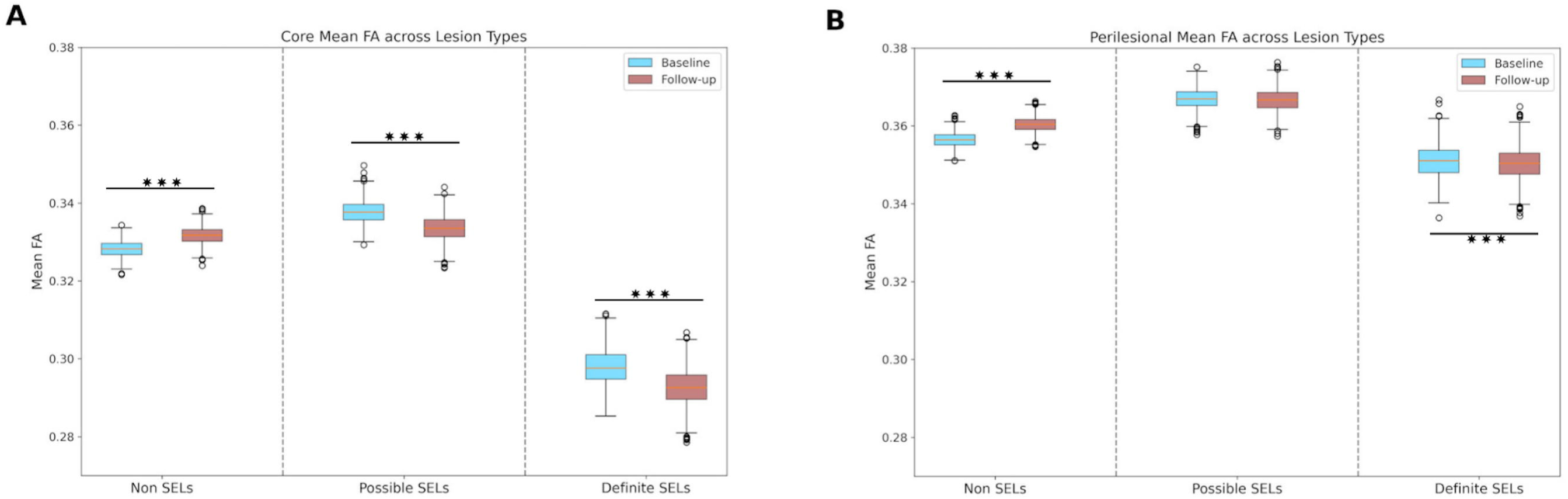
“FA distribution within core and perilesional areas across lesion types at baseline and last MRI”. Plots of the lesion core (panel A on the left) and the perilesional area (panel B on the right) distribution of bootstrap-computed mean FA values across lesion categories (i.e. definite SELs, possible SEL and non-SELs). For each lesion category, a pair of boxplots is presented: the left box represents the baseline data (in light blue), and the right box shows the last MRI follow-up (in red), both derived using bootstrap methods. *** indicate p<0.001

To assess whether lesion location influences microstructural integrity, we performed a one-way ANOVA on FA values across lesions in deep white matter (DWM), juxtacortical, and periventricular regions, revealing a significant effect of location (F = 32.16, *p*<0.001). Post-hoc analysis exploring spatial areas showed that DWM lesions had significantly higher FA than both juxtacortical and periventricular lesions (*p* < 0.001; Supplementary Figure 2).

Longitudinally, FA decreased in both the core and the perilesional area of definite SELs (β=-5.3 ×10□□, 95% CI −5.8 to −2.0, p<0.001; β=-3.8 ×10□□, 95% CI −6.5 to −0.7, p=0.006, respectively). The core and the perilesional FA within possible SELs remained stable over time. Non-SELs showed an opposite trend, with FA increasing in both areas (β=1.9 ×10□□, 95% CI 0.4 to 3.5, p=0.019; β=1.8 ×10□, 95% CI 0.5 to 3.2, p=0.008, respectively). The results for the other diffusivity metrics are reported in Supplementary Table 2.

### Correlations of FA metrics across lesion types with other MRI metrics

In the subject-level correlation analyses, annualised PBVC had a significant correlation with the mean FA longitudinal change within both the core and the perilesional areas of definite SELs (ρ=0.27, p=0.003; ρ=0.22, p=0.029, respectively). There was a slightly lower correlation with mean FA change in both the spatial areas of possible SELs (ρ=0.20, p=0.040; ρ=0.21, p=0.014, respectively), while no significant correlations in such metrics were found within non-SELs. We found a weak correlation between subject-level baseline lesion volume and mean core FA at baseline in definite SELs (ρ=-0.23, p=0.020, Supplementary Table 3). A moderate positive correlation was observed between mean NAWM FA and both core (_ρ_=0.50, p<0.001) and perilesional FA (ρ=0.61, p<0.001) in non-SELs at baseline. In contrast, correlations between NAWM and baseline FA in definite SELs were weaker, with a significant association only for perilesional FA (ρ=0.26, p<0.001), and no significant correlation within the core.

### Association of FA changes in lesion types with long-term clinical outcomes

Mixed-effects models evaluating the impact of FA changes in definite SELs on long-term clinical scores are summarised in Table 2. FA decline in both the core and perilesional area of definite SELs was significantly associated with reduced MSFC over time and all the z-score subcomponents (9HPT, T25FW, PASAT): as FA decreased, corresponding z-score changes in those metrics also dropped. The perilesional FA change in definite SELs was also significantly linked to SDMT. In pwMS with PIRA, the baseline core FA in definite SELs was associated with greater SDMT reduction (r=0.56, p=0.038) compared to the Stable group (r=0.15, p=0.345, Supplementary Figure 3). FA decrease in possible SELs (core and perilesional area) was associated with a reduction in PASAT and SDMT z-scores (Supplementary Table 4).

**Table 2.** Mixed-effects models assessing the clinical outcomes predicted by SEL signature.

| Clinical measure | FA change in definite SEL (n=400)<br>Beta coeff., 95% CI, P value* |  |
| --- | --- | --- |
|  | Lesion Core | Perilesional Area |
| MSFC z-score change | 0.03 (0.01; 0.06),<br>**p=0.004** | 0.03 (0.01; 0.05),<br>**p=0.009** |
| 9HPT z-score change | 0.08 (0.02; 0.14),<br>**p=0.009** | 0.08 (0.02; 0.14),<br>**p=0.011** |
| T25FW z-score change | 0.02 (0.01; 0.04),<br>**p=0.005** | 0.03 (0.01; 0.04),<br>**p<0.001** |
| PASAT z-score change | 0.04 (0.01; 0.07),<br>**p=0.003** | 0.05 (0.02; 0.08),<br>**p<0.001** |
| SDMT z-score change | 0.04 (-0.01; 0.07),<br>p=0.057 | 0.04 (0.01; 0.08),<br>**p=0.043** |

\*Mixed-effects models using as the outcome the clinical variable and as a the predictor the change in FA (interaction term with time) in the different spatial localisation (lesion core and perilesional area), including as fixed effects the demographic variables (age, gender, disease duration), baseline lesion volume, PBVC and using as random effects the lesion and the subject-level identifiers, and the scanner protocol. All the models were applied to the subset of definite SELs (n= 400 lesions).
Abbreviations: Multiple Sclerosis Functional Composite = MSFC; Nine Hole Peg Test = 9HPT, Timed 25 Foot Walk = T25FW, Symbol Digit Modalities Test = SDMT, Paced Auditory Serial Addition Test = PASAT.

### Association of FA metrics in lesion types with PIRA

In a simple logistic model adjusted for age, baseline lesion volume, Jacobian expansion, lesion count, PBVC and baseline EDSS at the subject level, mean core FA in definite SELs predicted PIRA risk (OR=0.01, 95% CI 0.01-0.12, p=0.001), while no significant association for perilesional FA in these lesion types was found. No significant associations were identified between core/perilesional FA in possible SELs or non-SELs and PIRA.

A Cox regression model to evaluate the time to reach PIRA at the patient level showed that a faster PIRA onset was significantly associated with lower mean FA values over time in the core and the perilesional area of definite SELs (HR=0.03 95% CI 0-0.49, p=0.015, HR=0.11 95% CI 0.01-0.95, p=0.045, respectively), higher Jacobian expansion, a higher count of definite SELs, larger PBVC decrease, higher baseline age and male gender. On the contrary, the baseline volume of definite SELs were not predictive of PIRA (Figure 3). Neither core/perilesional FA within possible SELs and non-SELs was associated with time to PIRA (Supplementary Table 5). A post-hoc analysis limited to patients with RRMS produced results consistent with those observed in the overall cohort (Supplementary Figure 4).

**Figure 3.**
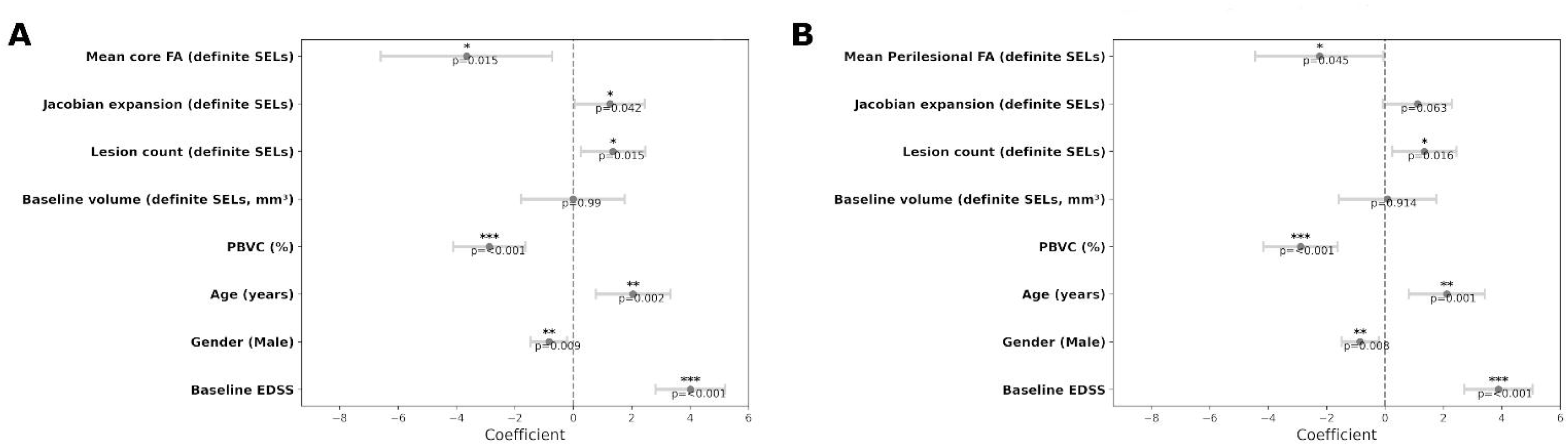
“Time-to-event analysis to assess the relationship between PIRA hazard and FA within the core and perilesional area of definite SELs”. Forest plots of the Cox’s multivariate model are described in the tables below, illustrating hazard ratios for time to reach PIRA, using SEL diffusivity, specifically mean FA from both core and perilesional areas. The left plot (A) displays results for the FA within the core of definite SELs, while the right plot (B) exhibits the FA within the perilesional area of definite SELs. In those models, the FA values were obtained over all MRI time points, and each model integrates age, gender, baseline EDSS, PBVC, Jacobian expansion, lesion count and baseline volume.

## Discussion

This study examined spatial structural MRI patterns in chronic active lesions by assessing diffusivity longitudinally within SELs in a cohort of pwMS with mild disease. This analysis demonstrated a distinctive FA signature in SELs, indicating significant microstructural progressive damage in both the core and perilesional areas of such lesions over time, which was associated with long-term MS physical and cognitive disability. Longitudinal FA measures within the core and perilesional areas of SELs predicted a higher risk and faster progression to PIRA.

In this cohort, 22% of subjects exhibited PIRA and had longer disease duration and older age compared to the clinically stable group, aligning with previous observations^33^. Moreover, pwMS with PIRA showed accelerated brain atrophy rates, as previously reported^34^, and a trend towards increased baseline SEL volume. Our findings suggest a significant association between imaging markers of chronic activity and PIRA. Similarly, elevated glial fibrillary acid protein (GFAP), as a blood-derived marker of MS progression, have demonstrated similar associations with PRLs^35^.

In the lesion-level analysis, definite SELs showed the lowest FA values: over about two years, both the core and the perilesional FA declined consistently with a prevalent decrease in the core indicating concentrated central tissue damage, while non-SELs had an opposite trend of increased FA. Thus, we hypothesise that such distinctive anisotropy signature in definite SELs likely represents pathological changes in chronic active lesions^6^, such as a persistent and constant alteration of the white matter fibre packing, possibly corresponding to severe myelin damage associated with chronic MS activity. Conversely, the observed FA increase within non-SELs could reflect ongoing remyelination processes.

Previous research also identified a subtype of extensively damaged white matter lesions at DTI^17^. Similarly, Elliott et al. demonstrated that SELs were distinguished by baseline lower FA and higher radial diffusivity (RD), with more pronounced longitudinal RD increases, particularly evident in progressive MS^36^. Recently, lower baseline myelin content and greater myelin loss over time were found in SELs at 7T MRI^37^. Our results expand this work through the exploration of a clinically available MRI measure of anisotropy within the core and the perilesional areas of chronic MS lesions and their association with long-term disability. To minimize the number of analyses and reduce the risk of type I error, we focused on FA measures; however, changes in the perilesional and core regions of SELs observed with other DTI metrics showed similar trends (Supplementary Table 2).

As a key finding, the altered FA signature in definite SELs was associated with other radiological markers of MS burden. Furthermore, a weak but positive correlation between brain atrophy measures and mean FA change, both in perilesional regions and the core of SELs, supports the hypothesis that SELs serve as markers of disease progression. Lastly, while NAWM FA did not correlate with core FA in definite SELs at baseline, a moderate correlation with FA in possible SELs and non-SELs suggests that the core of definite SELs undergoes distinct damage, whereas other lesion types and perilesional areas retain structural similarities to NAWM.

While SELs have not been extensively investigated in large cohorts using post-mortem brain samples, a recent study suggested a link between lesion expansion in MS and its chronic active state at pathology^38^. MRI *in vivo* analyses have confirmed short-term expansion in SELs over a timeframe of around 2 years^39^, while only a few long-term studies on SELs exist^40^ due to the inherent challenge of indefinitely expanding white matter lesions. It is presumed that SELs may reach a plateau in volume increase and potentially counterbalance atrophy accumulation due to gliosis. Recent multimodal MRI research found that lesions labelled as both SELs and PRLs exhibit greater microstructural damage, indicated by reduced FA and increased RD^18^. Despite modest correlations between PRLs and SELs and rare co-localization, their presence in the same patient may signal a more aggressive disease course^12^.

The findings of this study further underscore the predictive value of diffusivity signatures in SELs for long-term physical and cognitive disability, particularly among pwMS developing PIRA. Specifically, reductions in core and perilesional FA within definite SELs were significantly correlated with worsening in MSFC, 9HPT, and PASAT z-scores over approximately nine years, highlighting their potential as markers of MS progression. Importantly, these results are particularly relevant given that the cohort analysed exhibited only mild disability, emphasising the utility of these metrics in early disease stages. Ultimately, we found that the elevated odds of belonging to the PIRA group and higher hazard for a shorter time to the first onset of PIRA in the time-to-event analysis were predicted by FA reductions within the core and perilesional area of the definite SELs. Furthermore, the models pointed out that the Jacobian expansion and the count of SELs serve as predictors of an increased hazard for earlier onset of PIRA. This highlights the potential significance of dynamic lesion behaviour in disease progression, complementing measures of intrinsic microstructural alterations.

This study has limitations related to cohort size and diffusivity measurement methods. Firstly, the low number of pwMS with worsening disability reduced the rate of PIRA events, limiting statistical power. A potential limitation is the inclusion of progressive MS patients, which may bias results due to their higher risk of PIRA; however, the exclusion of these cases did not alter the significance of key findings, supporting the robustness of our results.

Additionally, the low specificity of DTI for MS pathology complicates interpretation due to structural complexities, such as crossing fibres. The implementation of DTI in clinical settings could incur significant additional costs and require adjustments to radiology protocols. Furthermore, SEL analysis can be influenced by image quality, registration methods, and lesion classification criteria, and the lack of standardised frameworks for SEL classification and diffusion MRI techniques in MS is due to the variability in acquisition sequences, each offering different specificity for chronic active lesion characteristics. Another limitation is the absence of spinal cord imaging and PRL data, as other relevant predictors associated with MS clinical progression. Lastly, MRI measures related to treatment effects were not examined due to cohort heterogeneity.

Future research should focus on the biological basis of SELs and their connection to chronic active lesions. Methodological studies are needed to refine SEL algorithms, determine optimal follow-up intervals for detecting lesion expansion, and establish a cut-off expansion rate for sensitivity analyses. Advanced quantitative MRI methods may replace conventional DTI, offering greater specificity for chronic active lesions. Improving techniques to quantify lesion expansion and integrate multi-modal MRI sequences could enhance the prediction of lesion evolution linked to PIRA.

In conclusion, pwMS have a distinct subtype of expanding chronic active lesions, detected as SELs, from diagnosis, marked by unique diffusivity features at their core and perilesional areas. These changes reflect ongoing microstructural damage contributing to disability progression. The decline in FA within SELs correlates with increased disability and faster progression to PIRA. Diffusivity SEL signatures are linked to higher disability and silent MS progression, offering potential as markers for risk stratification and guiding therapeutic strategies.

## Data Availability

Imaging data in BIDS format and the clinical data supporting the findings of this research can
be accessed upon reasonable request to the corresponding authors.

## Acknowledgements

We acknowledge the valuable contributions of all the physicians, psychologists, optometrists, nurses, and research coordinators involved in the research trial. Montserrat Artola, Ana Hernando, and Silvia Alvarez contributed by supporting the clinical aspects of the study visits as part of the nursing team. Laura Planas served as the research trial coordinator and was responsible for organising the study visits. The Centre de Diagnòstic per la Imatge (CDI) at the Hospital Clínic de Barcelona and the Imaging Platform at IDIBAPS provided essential collaboration in performing and processing the MRI scans. The authors are especially grateful to all study participants and their families for their willingness to participate and provide consent for this observational study.

## Declaration of conflicting interests

A. Calvi is supported by the Joan Rodés-Josep Baselga Advanced Research Contract from the fundación BBVA − Hospital Clinic Barcelona (2023), previously received the European Committee on Treatment and Research in MS (ECTRIMS) post-doc fellowship (2022), UK MS Society PhD studentship (2020). He received reimbursements for attending symposiums by ECTRIMS and North American Imaging in MS (NAIMS), Merck, Janssen, and has received consulting honoraria from QMENTA.

E. Martinez-Heras, F. Vivó, S. Alba-Arbalat have nothing to disclose.

E. Solana and E. Lopez-Soley received travel reimbursement from Sanofi and ECTRIMS.

E. Fonseca received funding for the ECTRIMS Clinical Training Fellowship Programme 2021-2022, speaking honoraria from Sanofi and travel reimbursement from UCB for national and international meetings.

M. Sepulveda received speaking honoraria from Roche and Biogen, and travel reimbursement from Roche, Biogen, and Sanofi for national and international meetings.

E. Martinez-Hernández received compensation for consulting services and speaker honoraria from Biogen, UCB, Argenx, and Alexion.

F. Prados received a Guarantors of Brain fellowship 2017-2020. F. Prados and B. Kanber are supported by National Institute for Health Research (NIHR), Biomedical Research Centre initiative at University College London Hospitals (UCLH).

A. Saiz received compensation for consulting services and speaker honoraria from Merck, Biogen-Idec, Sanofi-Novartis, Roche, Janssen, and Horizon Therapeutics.

Y. Blanco received speaking honoraria from Biogen, Novartis, and Genzyme.

P. Villoslada has stocks and has received consultancy payments from Bionure, Accure Therapeutics, Spiral Therapeutics, Attune Neurosciences, Adhera Health, Oculis, CLight, and NeuroPrex, none of them related to this article. He has also received consulting services and speaker honoraria from Eisai, Roche and BMS.

S Llufriu received compensation for consulting services and speaker honoraria from Biogen Idec, Novartis, Bristol Myer Squibb, Sanofi, Johnson & Johnson, and Merck. She holds grants from the ISCIII (PI21/01189), AGAUR (021-SGR-01325), and research support from Bristol Myers Squibb.

## Contributorship statement

The research project was conducted by Dr. A. Calvi, who was responsible for the study design, data collection, and statistical analysis. F. Vivo, E. Martinez-Heras, and E. Solana provided support and guidance during the data processing analysis and statistical evaluation phase. F. Prados and B. Kanber were involved in image pipelines and computational support. S. Alba-Arbalat, E. Lopez-Soley, Dr M. Sepulveda, Dr E. Martinez-Hernández, Dr J.M. Cabrera, Dr E. Fonseca, Dr S. Medrano, Dr A. Saiz, Dr. Y. Blanco, and Dr. P. Villoslada contributed to the study by participating in the data collection phase, providing and caring for study patients. Dr S. Llufriu served as the guarantor of the study.

