## Supplementary Tables and Figures for "Diffusivity anisotropy signature of the slowly expanding lesions predicts progression independent of relapse activity in multiple sclerosis"

**Supplementary Table 1**. Specifications of the MRI sequences

| SCANNER | SEQUENCES | REPETITION TIME (TR), MS | ECHO TIME (TE), MS | INVERSION RECOVERY TIME (TI), MS | NUMBER OF SLICES (N) | VOXEL SIZE  (MM) | DIFFUSION GRADIENTS AND B-VALUE(S) |
| --- | --- | --- | --- | --- | --- | --- | --- |
| 3T Magnetom Trio  (n = 64)  (n = 51) | MPRAGE    FLAIR    DWI:     (2) DTI      (2) HARDI | 1800    5000        6900    12600 | 3.01    379        89    112 | 900    1800        -    - | 240    208        55    80 | 0.94    0.94        2.5    2 | -    -        30 at 1000 s/mm^2^    60 at 1500 s/mm^2^ |
| 3T Magnetom Prisma fit  (n = 15) | MPRAGE    FLAIR    DWI  (PA and AP) | 1800    5000    5400 | 3.01    379    113 | 900    1800    - | 240    208    100 | 0.94    0.94    1.5 | -    -    30 at 1000 s/mm^2^  60 at 2000 s/mm^2^  90 at 3000 s/mm^2^ |

Abbreviations: MPRAGE = Magnetization-Prepared Rapid-Gradient-Echo, FLAIR = Fluid Attenuated Inversion Recovery, HARDI = High Angular Diffusion Imaging

**Supplementary Table 2**. Longitudinal changes of diffusivity (FA, RD, MD, AD) within the core and perilesional area grouped by subtype of lesions

| **Spatial localisation** | **Lesion type** | **FA change*** | **RD change*** | **MD change*** | **AD change*** |
| --- | --- | --- | --- | --- | --- |
| Lesion core | Non-SELs (n=3020) | 1.9 ×10^-4^ (0.4, 3.5), **p=0.0186** | -2.4 ×10^-4^  (-3.2, -1.6), **p=0.001** | -2.4 ×10^-4^  (-3.1, -1.7), **p<0.001** | -2.3 ×10^-4^  (-3.1, -1.6), **p<0.001** |
|  | Possible SELs (n=1391) | -1.2 ×10^-4^ (-3.3, 1.2), p=0.297 | 2.2 ×10^-4^  (0.8, 3.6), **p=0.002** | 2.7 ×10^-4^  (1.4, 4.0), **p<0.001** | 4.0 ×10^-4^  (2.6, 5.4), **p<0.001** |
|  | Definite SELs (n=400) | -5.3 ×10^-4^ (-5.8, -2.0), **p<0.001** | 2.0 ×10^-4^  (0.2, 3.8), **p=0.024** | 2.4 ×10^-4^  (0.6, 4.1), **p<0.001** | 3.7 ×10^-4^  (2.1, 5.3), **p<0.001** |
| Perilesional area | Non-SELs (n=3020) | 1.8 ×10^-4^ (0.5, 3.2), **p=0.008** | -0.9 ×10^-4^  (-1.7, -0.3), **p=0.007** | -0.3 ×10^-4^  (-0.9, 0.3), p=0.3556 | 1.3 ×10^-4^  (0.5, 2.0), **p<0.001** |
|  | Possible SELs (n=3020) | -0.9 ×10^-4^ (-2.9, 1.2), p= 0.325 | 2.0 ×10^-4^  (0.8, 3.1), **p=0.001** | 2.2 ×10^-4^  (1.1, 3.4), **p<0.001** | 3.2 ×10^-4^  (1.9, 4.4), **p<0.001** |
|  | Definite SELs (n=400) | -3.8 ×10^-4^ (-6.5, -0.7), **p=0.006** | 2.3 ×10^-4^  (0.9, 3.8), **p=0.001** | 2.6 ×10^-4^  (1.2, 3.9), **p<0.001** | 2.9 ×10^-4^  (1.4, 3.9), **p<0.001** |

^*^Beta coefficient (95% confidence intervals), p-value relative to the mixed-effects models that evaluate the diffusivity change (FA, RD, MD and AD) as the outcome and the predictor variable time from baseline MRI scan (in months), after adjusting for normalised lesion volume, PBVC and using as random effects the patient and lesion level unique identification numbers and the scanner protocol. The models were run at the lesion level within each volumetric subtype. In bold are the significant differences, set as a p-value <0.05.

Abbreviations: FA=fractional anisotropy, RD = radial diffusivity, MD = mean diffusivity, AD = axial diffusivity

**Supplementary Table 3.** Association between core and edge diffusion metrics (at baseline) by lesion types and other MRI metrics

| Correlation matrix variables |  | **Baseline FA metrics (mean) by lesion type and spatial area**  Correlation coefficients (ρ Spearman, p-value) | | | | | |
| --- | --- | --- | --- | --- | --- | --- | --- |
|  | Lesion type | Definite SEL | | Possible SEL | | Non-SEL | |
|  | Spatial area | Core | Perilesional | Core | Perilesional | Core | Perilesional |
| **Baseline global lesion volume (mean)** |  | **-0.23, p=0.020** | -0.04, p=0.720 | -0.02, p=0.830 | 0.13, p= 0.150 | 0.04, p=0.610 | 0.14, p=0.120 |
| **PBVC annualised**  **(%)** |  | 0.10, p=0.300 | 0.03, p=0.750 | 0.13, p=0.150 | 0.07, p=0.450 | **0.17, p=0.050** | **0.20, p=0.020** |
| **NAWM FA**  **(mean)** |  | 0.07, p=0.220 | **0.26, p<0.001** | **0.24, p<0.001** | **0.43, p<0.001** | **0.50, p<0.001** | **0.61, p<0.001** |

Correlation coefficients were obtained as Spearman´s rho between the patient-level FA metrics divided by type after Combat harmonisation and adjusting for sex and age, and the patient-level mean lesion volume (total sum of all lesion types) or the mean NAWM FA. In bold indicated the correlations with significance level p<0.05.

Abbreviations: NAWM= normal-appearing white matter, PBVC = percentage brain volume change, FA = fractional anisotropy.

**Supplementary Table 4.** Mixed-effects models assessing the clinical outcomes predicted by FA change within possible SELs and non-SELs

| **Clinical measure** | **FA change in possible SELs**  (n=1391)  Beta coeff., 95% CI, P value**^*^** | | **FA change in non-SELs**  (n=3020)  Beta coeff., 95% CI, P value**^*^** | |
| --- | --- | --- | --- | --- |
|  | Lesion Core | Perilesional Area | Lesion Core | Perilesional Area |
| MSFC z-score change | 0.01 (-0.01; 0.01), p=0.634 | -0.01 (-0.01; 0.01), p=0.986 | -0.01 (-0.01; 0.01), p=0.220 | -0.01 (-0.01; 0.01), p=0.522 |
| 9HPT z-score change | 0.01 (-0.01; 0.01), p=0.401 | 0.01 (-0.01; 0.03), p=0.401 | -0.01 (-0.01; 0.01), p=0.402 | -0.01 (-0.01; 0.01), p=0.402 |
| T25FW z-score change | -0.01 (-0.01; 0.01), p=0.629 | -0.02 (-0.01; 0.01), p=0.629 | 0.01 (-0.01; 0.01), p=0.067 | 0.01 (-0.01; 0.01), p=0.068 |
| PASAT z-score change | 0.02 (0.01; 0.03), **p=0.002** | 0.03 (0.01; 0.04), **p<0.001** | 0.01 (-0.01; 0.01), p=0.465 | 0.01 (0.01; 0.02), **p=0.010** |
| SDMT z-score change | -0.01 (-0.03; -0.01), **p=0.018** | -0.02 (-0.03; -0.01), **p=0.023** | -0.01 (-0.01; 0.01), p=0.265 | -0.01 (-0.01; 0.01), p=0.380 |

^*^Mixed-effects models using as the outcome the clinical variable and as the predictor the change in FA (interaction term with time) in the different spatial localisation (lesion core and perilesional area), including as fixed effects the demographic variables (age, gender), normalised lesion volume, PBVC and using as random effects the lesion and the subject-level identifiers, and the scanner protocol. All the models were applied to the subset of possible SELs (n=1391 lesions) and non-SELs (n=3020).

Abbreviations: Multiple Sclerosis Functional Composite = MSFC; Nine Hole Peg Test = 9HPT, Timed 25 Foot Walk = T25FW, Symbol Digit Modalities Test = SDMT, Paced Auditory Serial Addition Test = PASAT.

**Supplementary Table 5.** Cox regression models evaluating time to reach PIRA and FA within possible SELs and non-SELs

| Model | Variables | Time to PIRA  HR (95% CI), p-value | Model | Variables | Time to PIRA  HR (95% CI), p-value |
| --- | --- | --- | --- | --- | --- |
| possible SELs -core | Core FA (possible SELs) | 0.26 (0.05, 1.42), p=0.12 | non-SELs - core | Core FA (non-SELs) | 2.78 (0.37,21.14), p=0.323 |
|  | Jacobian expansion (possible SELs) | 51.44 (9.39,281.7), **p<0.001** |  | Jacobian Expansion (non-SELs) | 0.01(0, 0.03), **p<0.001** |
|  | Lesion count (possible SELs) | 2.71 (0,4,18.27), p=0.306 |  | Lesion count (non-SELs) | 7.31 (1.18,45.08), **p=0.032** |
|  | Baseline volume (possible SELs) | 0.68 (0.07, 7.09), p=0.747 |  | Baseline volume (non-SELs) | 1.44 (0.39, 5.35), p=0.59 |
|  | PBVC | 0.04 (0.01, 0.22), **p<0.001** |  | PBVC | *Deleted for high multicollinearity* |
|  | Age | 18.92 (3.87, 92.6), **p<0.001** |  | Age | 26.6 (5.5, 128.57), **p<0.001** |
|  | Gender (male) | 0.47 (0.19,1.17), p=0.104 |  | Gender (male) | 0.43 (0.18, 1.06), p=0.067 |
|  | EDSS (baseline) | 6.8 (0.88, 52.67), p=0.067 |  | EDSS (baseline) | 1.76 (0.3, 10.5), p=0.503 |
| possible SELs - perilesional | Perilesional FA (possible SELs) | 0.26 (0.04, 1.54), p=0.138 | non-SELs - perilesional | Perilesional FA (non-SELs) | 1.87 (0.19, 18.83), p=0.595 |
|  | Jacobian expansion (possible SELs) | 60.31 (10.75,338.2), **p<0.001** |  | Jacobian expansion (non-SELs) | 0.01 (0.0,0.02), **p<0.001** |
|  | Lesion number (possible SELs) | 2.49 (0.37, 16.67), p=0.347 |  | Lesion number (non-SELs) | 6.58 (1,43.86), p=0.051 |
|  | Baseline volume (possible SELs) | 0.65 (0.06, 6.5), p=0.713 |  | Baseline volume (non-SELs) | 1.27 (0.35, 4.66), p=0.719 |
|  | PBVC | 0.04 (0.01, 0.23), **p<0.001** |  | PBVC | *Deleted for high multicollinearity* |
|  | Age | 21.05 (4.32, 102.6), **p<0.001** |  | Age | 27.6 (5.71, 133.6), **p<0.001** |
|  | Gender (male) | 0.46 (0.18, 1.17), p=0.102 |  | Gender (male) | 0.43 (0.17, 1.04), p=0.062 |
|  | EDSS (baseline) | 6.38 (0.83, 49.03), p=0.075 |  | EDSS (baseline) | 1.67 (0.28, 9.82), p=0.57 |

Cox regression models evaluate the event time to reach PIRA, predicted by the core FA (upper) and perilesional FA (lower) values within the possible SELs and non-SELs (left and right), and the other covariates.

**Supplementary Figure 1.** Effect of ComBat harmonization

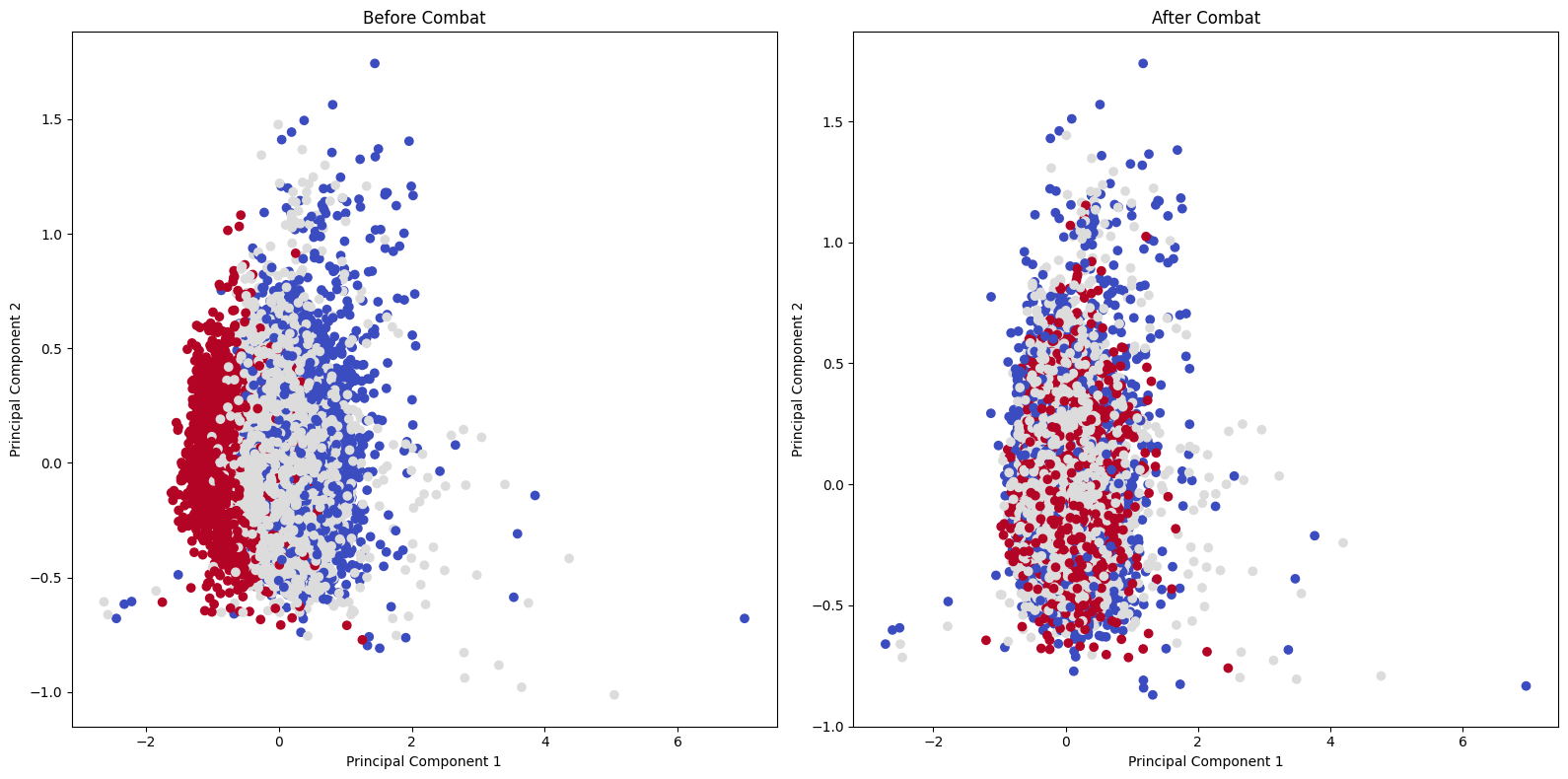

The data shows distinct clustering by the scanner protocol, indicating variability introduced by different DWI protocols in the left panel (before ComBat harmonisation). The clustering by the protocol used is notably diminished after ComBat (right panel after ComBat, ANOVA p<0.001), though some residual effects remain, as illustrated in the accompanying principal component analysis (PCA) plots.

Red colour indicates the first scanner DWI protocol, grey colour indicates the second DWI protocol and blue colour indicates the third protocol.

**Supplementary Figure 2.** Lesion-level FA values across spatial brain areas.

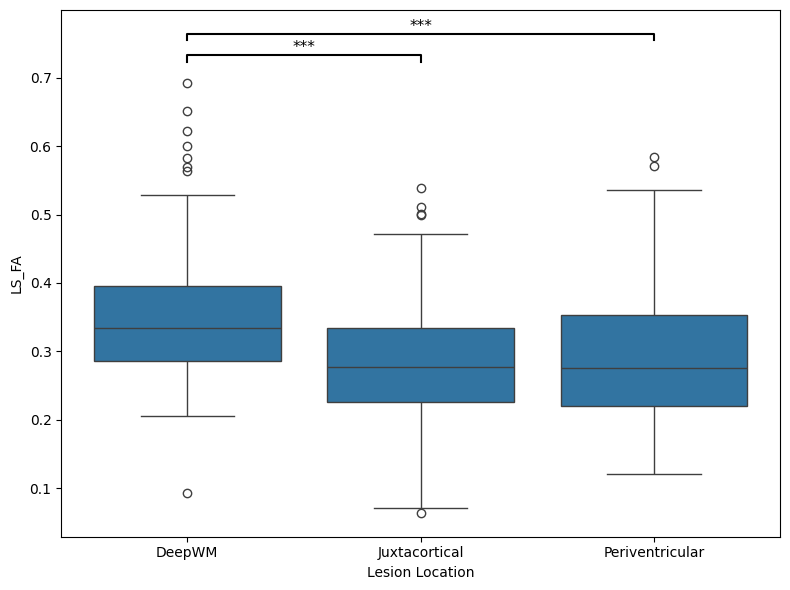

Boxplots showing the results of lesion-level FA (LS_FA) across Deep White Matter (DeepWM), Juxtacortical, and Periventricular spatial area. The ANOVA test showed a highly significant effect for lesion location (F=32, 16 p<0.001). Lesions in Deep WM exhibit significantly higher FA than Juxtacortical and Periventricular lesions after post-hoc comparisons (Tukey´s HSD, p<0.001). No significant difference was found between Juxtacortical and Periventricular lesions.

**Supplementary Figure 3.** Association between baseline FA within definite SELs and SDMT, grouped by PIRA versus stable

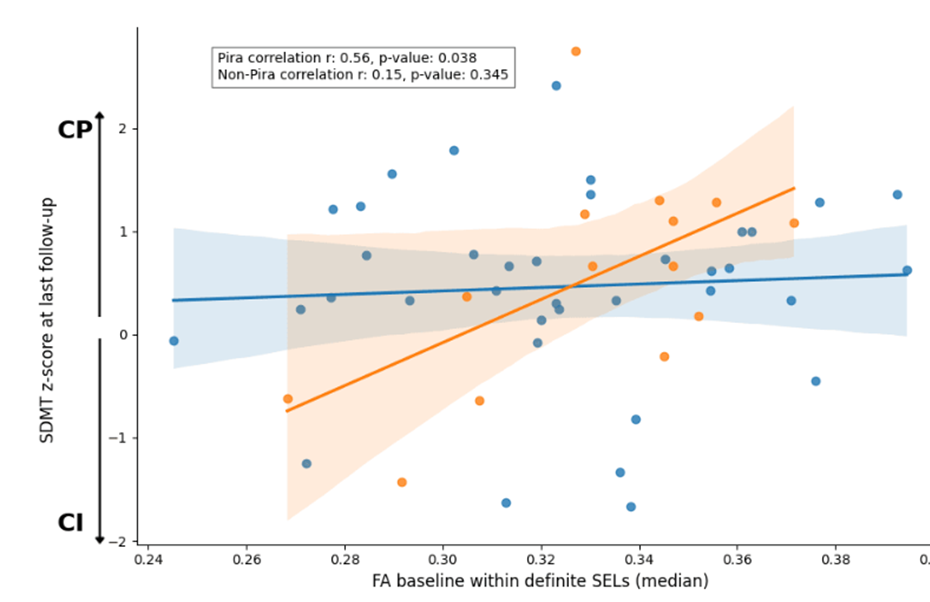

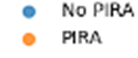

Stable

PIRA

Scatter plot presenting the association of the subject level mean values of FA (at baseline) within definite SELs and the SDMT z-score (at last follow-up assessment) in pwMS who developed PIRA (r=0.56, p=0.038) compared to the stable group (r=0.15, p=0.345). The blue dots indicate the group of pwMS who did not develop PIRA (stable), while the orange colour represents the ones who reached PIRA. The arrow on the y-axis indicates that an increase in the SDMT z-score indicates subjects who are cognitively preserved (CP) while the decrease in the z-score indicates the subjects who are cognitively impaired (CI).

**Supplementary Figure 4.** “Time-to-event analysis to assess the relationship between PIRA hazard and FA within the core and perilesional area of definite SELs after excluding subjects with progressive MS phenotypes”

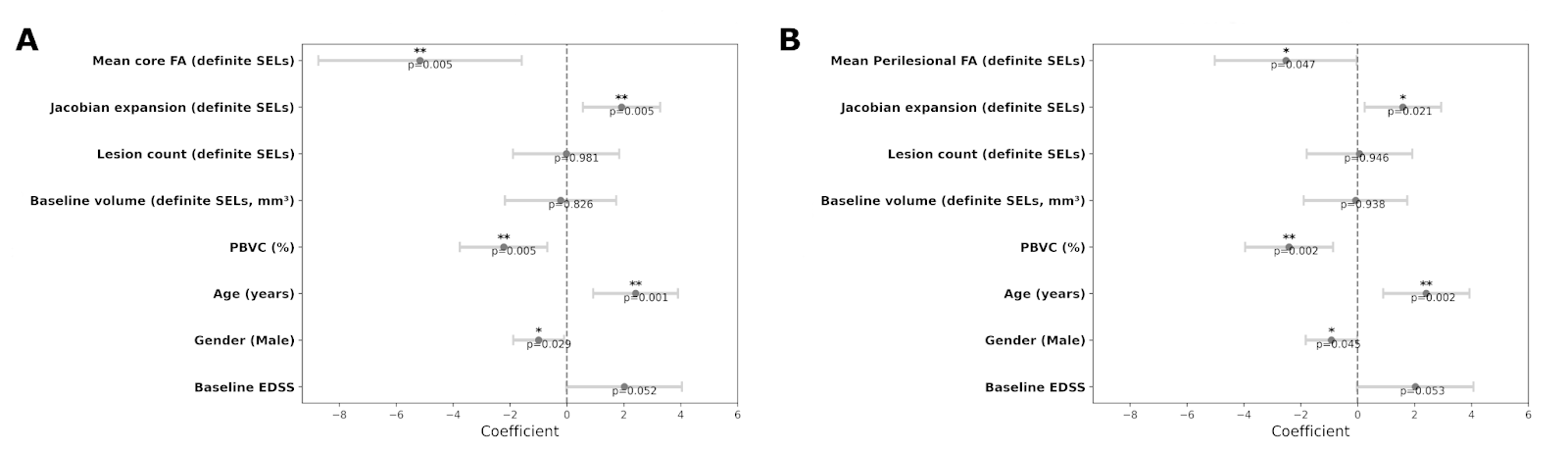

Forest plots of the Cox’s multivariate model are described in the tables after excluding pwMS with progressive phenotypes (either primary and secondary progressive MS), and hazard ratios for time to reach PIRA are presented, using SEL diffusivity, specifically FA, as predictors. Among 117 total, 17 subjects reached PIRA. The left plot (A) displays results for the FA within the core of definite SELs, while the right plot (B) exhibits the FA within the perilesional area of definite SELs. In those models, the FA values were obtained over all MRI time points, and each model integrates other relevant markers for PIRA, including age, gender, baseline EDSS, PBVC, lesion count, mean Jacobian expansion and definite SEL volume.

Abbreviations=FA: fractional anisotropy
